# Audit of Impact of a Clinician & Patient Support Tool in a Nurse-led Clinic for Inflammatory Bowel Disease Service at Milton Keynes University Hospital

**DOI:** 10.1101/2024.02.17.23300358

**Authors:** L Lewis, C Michie, A Sheppard, M.A Stone, G MacFaul, J Harrison

## Abstract

**Objective:** This audit examines the impact of a recently-commissioned clinician and patient support tool, on care pathways for Inflammatory Bowel Disease patients at Milton Keynes University Hospital, on follow up appointments (including PIFUs), associated cost savings and patient and clinician engagement and experience.

**Design:** The audit was conducted using a retrospective, consecutive case review of appointment records of patients enrolled onto the iOWNA platform during a 9-month audit period. Patient-clinician interactions were categorised depending on whether an appointment was saved and an appropriate nursing time saving logged. Data on both cost savings and PIFU numbers were also collected from the trust. Quantitative and qualitative feedback of clinician and patient experience were captured.

**Results:** The deployment of iOWNA resulted in a total time saving of 14,735 minutes across the audit period, of which 9,280 minutes comprised savings on in-person appointments, representing 232 appointments saved. The cost savings for these 232 replaced appointments totalled 16,704 GBP. The audit period also saw a statistically significant (p<0.01) increase in the number of Patient-Initiated Follow Ups (PIFUs). Patient and clinician surveys reflected positive experiences of the new care pathway among all service users.

**Conclusion:** The results of the audit demonstrate the important role digital tools can play in transforming existing care pathways to deal with the widespread challenges facing the NHS following the COVID-19 pandemic, with annual savings of over 345 million GBP for the NHS if the appointment cost savings demonstrated in this audit were replicated alongside a 5% reduction nationally in outpatient attendances.

**KEY MESSAGES:** *What is already known on the topic:*

- The deployment of evidenced-based digitally-enabled care pathways for Inflammatory Bowel Disease (IBD) is scarce. Traditional pathways are maintained, with patient education during appointments being time-consuming for clinicians and ineffective for patients, resulting in patients forgetting large amounts of what has been communicated to them and not being able to self-manage their condition optimally.
- Leveraging technology has been acknowledged as an important tool for patient empowerment thus aiding patient self-management, which will play an important role in the recovery of elective care services in the wake of the disruption caused by the COVID-19 pandemic.

*What this study adds:*

- We report the results of a clinical audit on the impact of a commissioned clinician and patient support tool, iOWNA, on existing IBD care pathways at Milton Keynes University Hospital 2023.

*How this may affect research, practice or policy:*

- The clinical evidence from the audit demonstrates that digital transformation of chronic disease care pathways is an effective means of improving the delivery of outpatient care, offering advantages in terms of cost, clinician experience, and patient experience.

## INTRODUCTION

### The context for digital transformation

The digital transformation of patient care pathways promises to revolutionise the efficiency of healthcare delivery and offers a multiplicity of benefits for patients, clinicians, and healthcare systems.^1^ Indeed, delivering a digital transformation of health services occupies a whole chapter in the NHS’s Long-Term Plan of healthcare priorities for the next decade, and continues to be a core focus of the health service’s priorities in the most recently published operational planning guidance.^2 3^ The impetus for the digitalisation of healthcare services has recently intensified in the wake of the COVID-19 pandemic, which has seen a paradigm shift towards a society-wide adoption of digital technologies to enhance existing workflows.^4^ This impetus is all the more pressing given the unprecedented challenges facing healthcare providers and service users post-pandemic, with the number of patients on the elective care backlog standing at 7.75 million in August 2023, an increase of 70% when compared with February 2020.^5^ Clearing this backlog requires novel and innovative approaches to outpatient care, with the adoption of digital patient support tools offering enormous potential for health services operating in a post-pandemic environment.

### The context for IBD and MKUH

Chronic diseases present an enormous challenge to population health and health services, affecting roughly 15 million people living in the United Kingdom and requiring intermittent treatment across prolonged periods of time.^6^ Inflammatory Bowel Disease (IBD) is one such chronic disease affecting the bowel characterised by relapses and remission, which requires ongoing patient management, with an estimated 347,973 patients using IBD services in the UK.^7^ Streamlining IBD care pathways is essential for ensuring that these patients receive the correct treatment for their individual needs in the fastest possible time.

The need to implement a digital transformation of patient care pathways has been recognised by nurses in IBD clinics at Milton Keynes University Hospital (MKUH), where out-dated patient care pathways have put a strain on nurses, patients, and hospital budgets. This is a challenge which has been compounded by the structural inefficiencies ingrained in the still-dominant appointment-first model embarked upon by the NHS at its outset.^8^ In particular, unnecessary outpatient attendance for routine educational appointments and follow-ups are costly to the hospital and can be inconvenient for patients, while the manual distribution of information to patients in hard form through individual emails leads to a high volume of additional phone calls to clinics as patients struggle to self-manage their conditions. IBD as a specialty have championed patient initiated follow ups and was thus felt to be an ideal forum to deploy digital transformation with a clinician and patient support tool. A proven advanced digital clinician and patient support tool tailored for chronic diseases, iOWNA, has therefore been deployed to provide a digital solution to the aforementioned inefficiencies. iOWNA was commissioned for use in IBD clinics at MKUH with the backing of the hospital’s management having previously generated positive results in the rheumatology department, which the hospital hoped could be replicated across other chronic disease care pathways. The system enables clinicians working in chronic disease clinics such as IBD to send multimedia information directly to patients, and provides patients with a readily-accessible digital repository for this information so that they can better self-manage their condition without resorting to telephone helplines. Through enabling educational material and assessment questionnaires to be sent to patients, iOWNA also provides a digital-first option for educational and routine follow-up appointments within chronic care pathways.

## AIMS

The primary objective of this audit is to assess the impact of the implementation of iOWNA, a clinician and patient support tool, on the clinical pathways for IBD at MKUH. Specifically, the audit aims to provide a quantitative analysis of the efficiency savings derived from the implementation of iOWNA’s digital support tool in clinical practice for chronic disease, looking at both reductions in unnecessary in-person appointments and nursing time savings generated through the dissemination of materials to patients via iOWNA, along with data on iOWNA’s effect on patient-initiated follow-ups. Given that stakeholder engagement across a variety of levels plays a vital role in the successful development and implementation of systems for healthcare improvement, the audit also includes further quantitative, as well as qualitative, data collection from clinicians and patients on their experience of the transformation of IBD care pathways.^9^ The audit will then explore the wider implications of iOWNA’s impact on metrics which are fundamental for an effective healthcare service, and which broadly align with Bodenheimer’s expanded Quadruple Aim: reducing the cost of care, improving population health, enhancing the patient experience, and improving the experience of care staff.^10^

## METHODS

### Audit design

An audit was undertaken to evaluate the impact of the iOWNA system on care pathway efficiency in IBD clinics at MKUH. It is important to find a standard measurement for care pathway efficiency improvement outcomes as a primary endpoint when considering the adoption of digital health pathways. As such, the audit has leveraged data on nursing time savings from clinician-recorded records of patient interaction with the iOWNA support tool as a reliable metric for this purpose (these being exclusive of the administrative time required for the dissemination of material in either the previous or current care pathway).^11 12^ Quantitative analysis of clinic appointment data was then conducted over the first full nine months following the system’s implementation (1 February 2023 to 31 October 2023) to ensure the validity of results in the face of any seasonal variation. As secondary endpoints, patient and clinician feedback was collected towards the end of the audit period to measure the effects of the changed care pathways on service user experience.

### Procedure and Data Collection

Data collection was carried out for this audit by retrospective consecutive case review of patient appointment records from the IBD unit at MKUH pertaining to patients who had been on-boarded onto iOWNA in the audit period (n=422). Data was collected by nurses in a datasheet recording every instance of patient interaction with clinicians made through iOWNA. For each interaction, the following variables were recorded: the date of the interaction, and the type of content sent to the patient, and whether or not an appointment was saved.

➢ Interactions for which the iOWNA support tool replaced an in-person educational appointment when compared to previous patient care pathways were labelled on the data sheet as “YES”.
➢ Interactions for which the iOWNA support tool did not result in an in-person appointment replacement, but still contributed to time savings for the clinicians in comparison to previous care pathways due to the increased efficiencies in patient care, were labelled on the data sheet as “NO”.

For both of these categories, representative savings in nursing time were assigned as follows:

➢ For each of the interactions under the “YES” variable, an average saving of clinician time of 40 minutes was assigned, representing the length of the previous educational appointment with a nurse in an IBD clinic at the hospital.
➢ An approximation of time saving was applied for interactions under the “NO” variable given the range of communicative activities carried out through the iOWNA platform by the IBD nurses, and the difficulty in quantifying time-saving for every individual instance in a retrospective audit. An average saving of 20 minutes was assigned to the majority of patient-clinician interactions in the “NO” category, reflecting the approximate average nursing time saved dealing with patients in follow-up and helpline appointments conducted with the help of iOWNA, when compared to the standard length of nursing time allotted to such appointments in previous care pathways.
➢ For a small number of interactions under the “NO” variable (n= 4), a reduced average time saving was apportioned due to the nature of the interaction initiated by the clinician, for which a lower time saving was realised through the use of the iOWNA support tool.

Primary data collection was also carried out to ascertain whether any recorded impact on time savings made through the digital transformation of IBD patient care pathways was reflected by improvements in both clinician and patient experience.

On completion of the audit, surveys were sent to patients onboarded on the iOWNA patient care pathway. The surveys consisted of a series of 17 questions regarding their experience of the iOWNA platform compared to their experience of previous IBD care pathways, with questions written in consideration of the NHS 2023/24 priority that digital tools equip patients “to take greater control over their health and care”.^3^ Patient responses were recorded on a Likert scale, anchored at 0 and 10, in accordance with how far they agreed with/supported the statement in each question - ‘0’ representing total disagreement, ‘10’ total agreement.

Qualitative and quantitative data was also collected from every IBD nurse within the department (n=5) as a means of providing more extensive clinician stakeholder feedback of the redesigned digital care pathway. This took the form of anonymised questionnaires presenting a mixture of closed ‘yes/no’ questions along with limited space for explanations of answers, and a separate form inviting longer, descriptive responses related to their experience of workload and care pathways before and after the implementation of iOWNA.

Data on IBD patient initiated follow ups (PIFUs) was then collected from the hospital administration to determine whether the deployment of iOWNA had an impact on the rate of patients being placed on PIFU pathways.

### Data analysis

IBD patient records were stored in a structured dataset, double password-protected to ensure the security of patient data, which allowed for a thorough analysis of the data at the end of the audit period. Descriptive statistics were employed to assess the central tendencies and variability of the results, an important step in determining the applicability of the results to a broader context beyond the audit. The monthly mean and standard deviation (SD) for each measured variable were calculated to determine any trends or seasonal variations which might indicate the presence of external variables affecting the results. Cost savings were then calculated on the basis of the total number of appointments saved by the new clinic pathway multiplied by the cost of an IBD clinic appointment (£72 per new patient appointment).

The patient feedback surveys were then gathered into a single structured dataset and averages were collected from the quantitative feedback for each question, while a thematic analysis of the qualitative feedback received from IBD nursing team was conducted to identify recurring patterns in the clinician experience of the transformed patient care pathway.

### Ethical Approval

Due to the study taking the form of a retrospective clinical audit evaluating clinical practice, written patient consent and approval from a Research Ethics Committee were not required.

## RESULTS

The results of the analysis have been divided into two broad categories, each containing two subcategories. The results of the measurements of clinic efficiency savings have been broken down into clinician time saving, including appointment savings, and then cost savings for the hospital. User experience of the transformed care pathway has been subdivided into the quantitative analysis of patient experience and then the qualitative analysis of clinician experience.

### Clinic Efficiency Savings with iOWNA

Between 1 February and 31 October 2023, a total of 422 IBD patients had clinical interactions with IBD clinic nurses at MKUH, representing 31.7% of the 1331 active patients who were using IBD clinical services in the Hospital as of 10 October 2023.^13^ These 422 patients had a total of 506 interactions with clinicians through iOWNA in the audit period.

### Time savings

The deployment of the iOWNA’s digital support tool in clinical practice in IBD resulted in a total time saving of 14,735 minutes over the nine months of the audit period when compared to nurse time expended on previous care pathways [Figure 1]. Of this total, 9,280 minutes comprised savings on in-person clinic appointments (the “YES” variable on the datasheet), which were replaced by the distribution of educational material in multimedia form to patients through the platform. This represents 45.8% of all interactions carried out through iOWNA. Across the nine-month period, 232 clinic appointments were saved through the use of iOWNA’s digital support tool – an average of 25.8 appointments per month, SD = 9.08. The remaining 5,455 minutes represents the “NO” variable in the datasheet, that being time savings made to nursing activity through the use of iOWNA in which an appointment was not replaced.

**Figure 1.**
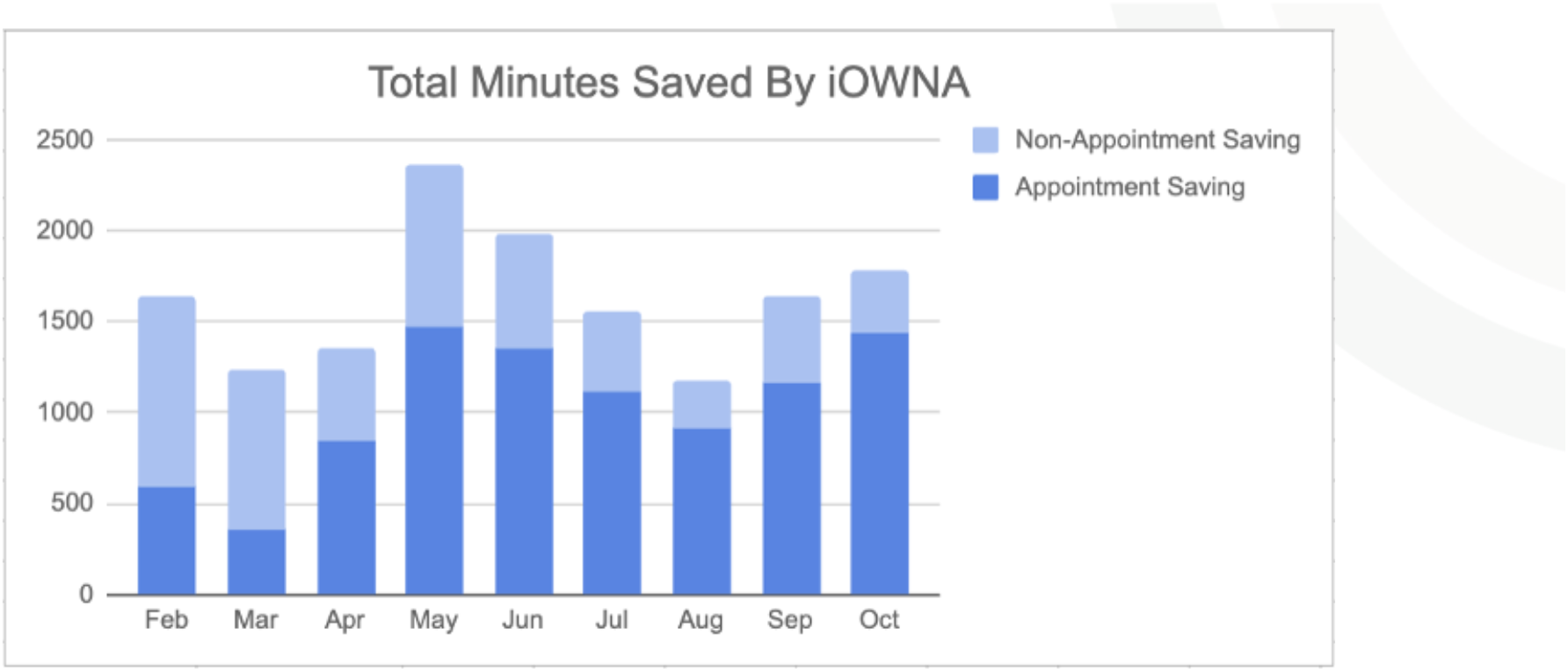
Total minutes of nursing time saved in new care pathway

Monthly variability in time saving was observed in the data, with March and August recording the lowest monthly time-saving, and May the highest. Monthly average timesaving = 1,637.22 minutes; SD = 351.18; with a coefficient of variation (CV) of 21.45%. Variability was more pronounced for appointment savings over the entire audit period (CV February to October = 35.21%), though was significantly less so after the first three months of the audit (CV May to October = 15.87%), potentially suggesting a period of adjustment to the implementation of the new digital care pathway [Figure 2]. The mild variability in appointment savings is also not unexpected given the variability of flare-up frequency among IBD patients.

**Figure 2.**
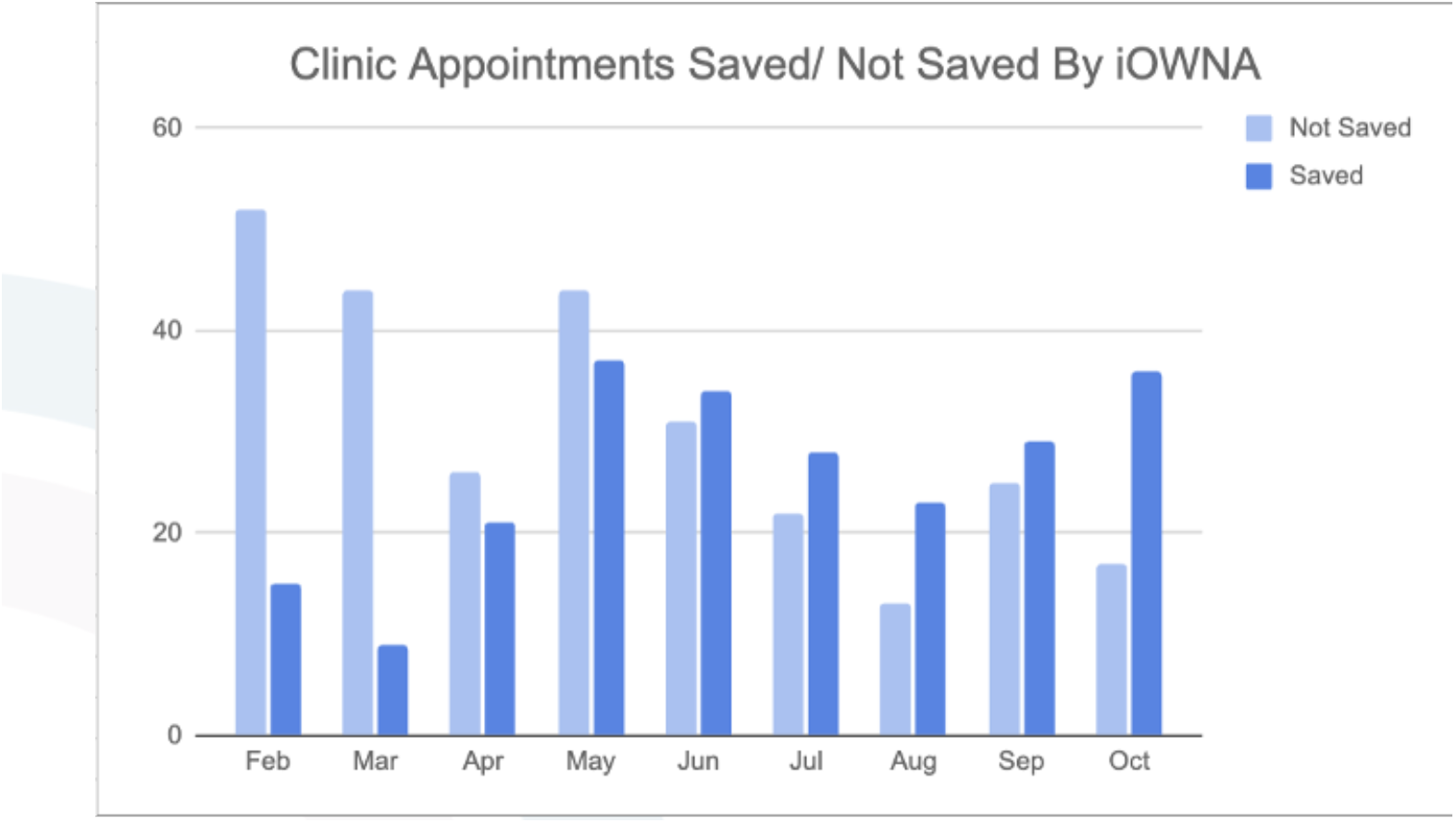
Clinical appointments which time savings through use of iOWNA each month, separated on the basis of whether a full appointment was replaced or not

### Cost savings

Cost savings for the 232 appointments which were replaced through iOWNA’s digital support tool amounted to a total of £16,704 across the audit period (£72 per appointment saved), with per capita savings of £39.58 per IBD patient using the system (n=422). Following this trend, a saving of £52,684 would be accrued in an equivalent nine-month period if all IBD patients at MKUH (n=1331) were to be on-boarded with iOWNA [Figure 3].

**Figure 3.**
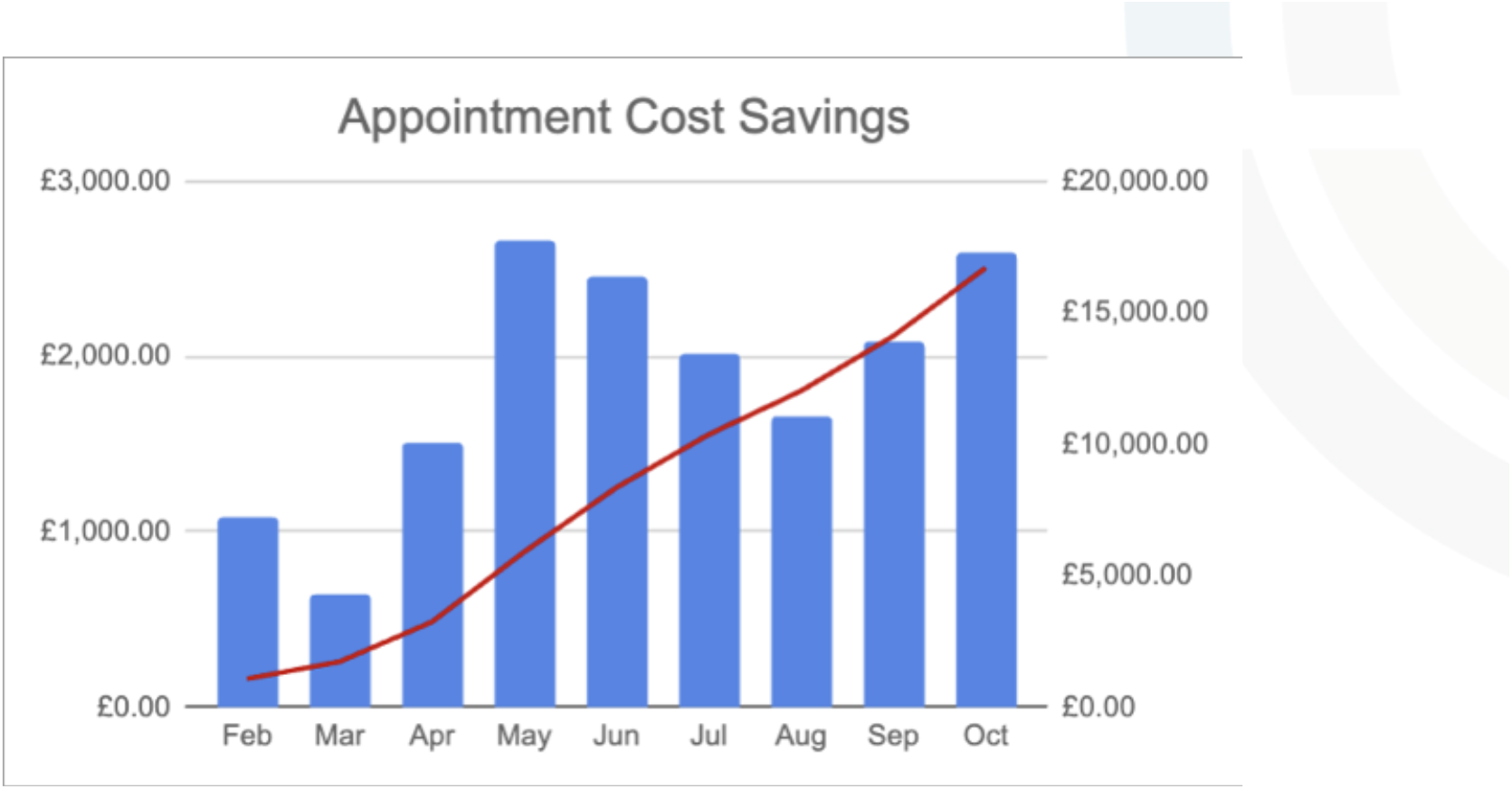
Cost savings through appointment replacement on the new care pathway, both monthly and cumulative

### Patient experience surveys

Average scores for the returned patient surveys (n=31), in which patients were invited to respond to statements about the platform’s utility with a score out of 10, gave a range of positive responses. The highest average (±SD) response was to the statement “usefulness of having information on device” (9.06±1.63), followed by “overall ease of iOWNA” and “ease of sign up and accessing iOWNA for the first time” (both 8.61±2.06). The lowest response was to the statement “reduced anxiety due to understanding treatment plan” (7.61±2.12), though this was still a notably positive result. An important metric of increased patient engagement came in the responses to the statement “improved patient experience and engaged more in treatment journey”, which saw a positive average response of 7.84±2.33.

### Staff feedback questionnaires

The responses to the anonymised feedback questionnaires (n=5) saw 100% of IBD nurses answer “yes” to the questions “do you feel iOWNA can benefit the IBD nurse/ patient”. The most common reasons ascribed to this benefit were savings in workload for nurses (n=4) and the ability for patients to better self-manage their care (n=3). The more descriptive responses to the second questionnaire, when organised thematically, likewise centred on iOWNA’s facilitation of patient self-management and reduction of nurses’ workload. The area of concern most commonly raised (n=3) with the adoption of iOWNA regarded the provision of materials to those who did not use a smartphone, an observation which should form the basis of a further research into the effects of age on the efficacy of digital care delivery.

### PIFU data

The data encompasses all IBD PIFUs in the six months prior to the audit period and the audit period itself (though the records for October are only partial, and so data analysis has only been conducted on records up to the end of September). The data reveals a notable increase in the number of PIFUs initiated during the audit period, with a monthly average of 64.63 from February to September 2023 as compared to the monthly average of 26 in the six months prior to the audit period [Figure 4]. A one-tailed T-Test was then conducted to determine the statistical significance of this increase, which returned a *p-*value <0.01, (*p=*0.00082).

**Figure 4.**
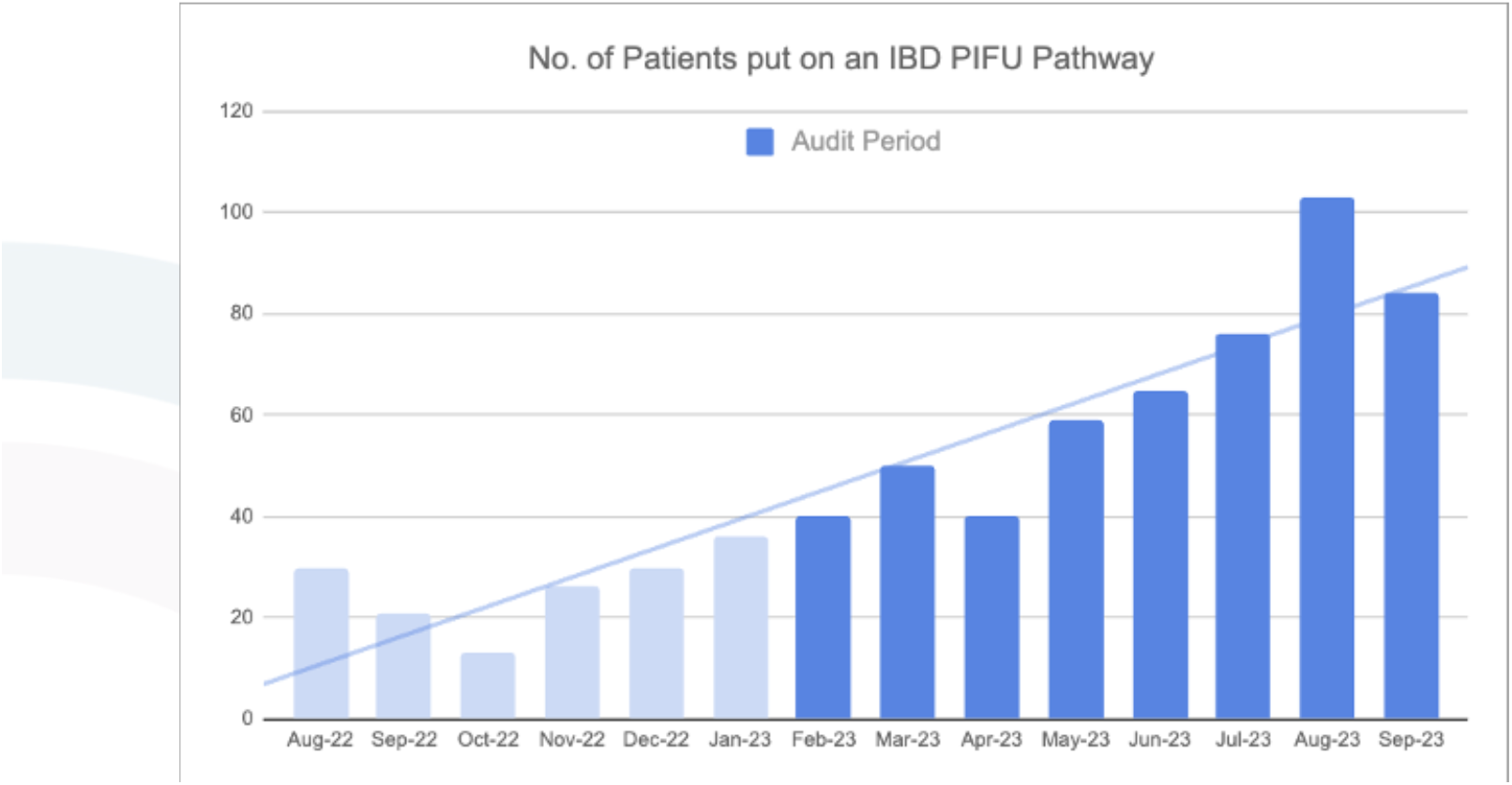
Number of IBD patients placed on PIFU pathways at MKUH

## DISCUSSION

The findings of this audit demonstrate the significant impact of the implementation of the iOWNA patient software system on outcome metrics which are meaningful for ensuring healthcare improvements that impact on each area of the Bodenheimer’s Quadruple Aim as well as on current national healthcare priorities.^10^

### Implications for healthcare efficiency and delivery

The implementation of a digital tool with the efficacy demonstrated by the iOWNA system has wide-ranging implications for improving healthcare efficiency and generating the step-change in care provision required to meet the current challenges of healthcare delivery. By reducing unnecessary in-person appointments and the amount of time spent administering information to patients through a digital-first system, healthcare organisations can allocate resources more effectively, seeing patients who are most in need of urgent care faster, leading to shorter waiting times.^14^ In consideration of the context of current public health policy, the digitally-enabled transformation of care pathway inefficiencies directly contributes to the NHS’s ambitious target of producing the annual efficiency gains of 2.2% in the healthcare system needed to recover core services in the wake of the COVID-19 pandemic.^15^

The clear correlation between the deployment of iOWNA and increased IBD PIFU rates is also significant in this regard, with the NHS’ 2023/24 Elective Care priorities setting out the transfer of patients onto PIFU pathways as an important part of their strategy to bring down the elective care backlog by reducing out-patient follow-ups by 25% against the pre-COVID (2019/20) baseline.^16^ Indeed, a recent study noted that adopting a national strategy of placing patients on digitised PIFU pathways would free up 1.4 million appointments annually.^17^ The increase can be attributed to iOWNA providing patients with the information required for self-management, which in turn has ensured that the IBD clinicians have had the confidence to place patients on PIFU pathways. This approach aligns with IBD UK’s national standards, which state that “patients should be supported in self-management”, as patient-activated pathways contribute to “better health outcomes, improved experiences of care and fewer unplanned care admissions” and are ideally suited to diseases such as IBD, which are characterised by often-unpredictable flare-ups.^18^

The financial implications of replacing in-person educational appointments with content distributed through a digital tool provide a further imperative for providing a digital-first offering for IBD outpatient appointments. Across the NHS in 2022/23 there were 95.9 million outpatient attendances – even a 5% reduction in this figure (well below the 33% envisaged in clause 1.48 of the NHS’s long term plan) would yield annual cost savings of over £575.4 million for the health service on the basis of a 2018 estimate of £120 per outpatient appointment, or £345.2 million on the conservative basis of the £72 cost of IBD appointments at MKUH.^19 20^ On a micro level, 45.8% of all interactions carried out through the platform in this audit resulted in an outpatient attendance saving. As this refers to the 31.7% of IBD patients using iOWNA at MKUH, the outpatient attendance reduction when measured proportionately against the entire IBD patient population of the hospital is 14.5%, a figure which would still have a significant financial impact if replicated on a wider scale.

A digitally-enabled transformation of care pathways must, however, at its centre be patient-focused, as efficiency gains cannot contribute towards improved health outcomes if they are achieved at the expense of patient safety or satisfaction. The structured feedback surveys from the audit provide quantitative confirmation of patient satisfaction with the redesigned IBD care pathway, as well as increased patient engagement with their treatment plan (with the statements “improved knowledge regarding condition” and “engaged more in treatment journey” eliciting the strongly positive responses of 8.16±2.08 and 7.84±2.33 respectively). This enhanced IBD patient engagement has been facilitated by the features of iOWNA’s support tool which enable clinicians to send information to patients in multimedia formats, including videos of instructions for patients of their treatment path, and aligns with the national healthcare priority to “put digital tools in place so patients can be supported with high quality information that equips them to take greater control over their health and care”.^3^ This shift to digitally-enabled care pathways also has widespread support among the patient population, with a recent report by the Health Foundation finding 78% of patients would be happy ‘to monitor their own health at home using technologies, instead of in a hospital’.^21^

For clinician service users, the ability of digital solutions to “optimise clinical processes to reduce the administrative burden” currently facing healthcare staff is recognised in section 5.16 of the NHS’s *North-Star*, its Long-Term Plan. 100% of the IBD nurses in the audit responded that iOWNA ‘can benefit the IBD nurse’, with the alleviation of workload a recurrent explanation for this answer. This alleviation of workload is the direct result of the replacement of care pathways which were characterised by the distribution of educational material through hard copy or email and high volumes of calls to IBD helplines from patients failing to manage their condition, with a tool enabling the digital storage and distribution of treatment information. Given that 34% of healthcare staff reported feeling burnt-out in the latest NHS staff survey, the roll-out of a streamlined digital care pathway as presented in this audit on a multi-departmental basis could have profound implications for staff satisfaction, and ultimately retention, across outpatient healthcare services.^22^

### Limitations

While this audit provides valuable insights into the impact of the iOWNA Patient Software System on clinic appointment savings at Milton Keynes Hospital, it is essential to acknowledge certain limitations. These can form the basis for further research into the efficacy of iOWNA’s patient-clinician support tool in clinical practice, which can inform future development to better meet the most pressing needs of patient care provision.

An acknowledged barrier to the uptake and efficacy of digital health tools is digital illiteracy, particularly among the elderly.^23^ This is reflected by current IBD clinical practice in MKUH, where digitally-enabled care pathways are not offered as standard for patients over 70, due to perceived issues surrounding the engagement of older patients with digital tools. The problem is notable though would appear to be fast receding as older generations become more technologically literate: while only 67% of over 65s reported using a smartphone in 2022 (the primary device for accessing iOWNA), compared to nearly 100% for the rest of the population, this a dramatic increase from the 3% of over 65s who reported using a smartphone in 2012.^24^ Conducting research into the intersection of age and efficacy of digital tools, and the possible means of mitigating any age-related access issues, would therefore be a useful future step in the wider deployment of iOWNA in clinical practice.

More broadly, digital health tools have shown promise as a means of mitigating and reducing systemic health inequalities, providing a means of improving access to healthcare among groups who have traditionally suffered from poor health outcomes.^25^ As such, it would also be beneficial in future studies into iOWNA’s efficacy to include demographic indicators as part of data collection in order to assess the impact of the platform’s deployment on certain groups within the population. Future studies should also quantify the carbon reduction savings associated with the reduction in patient journeys within digitally-enabled care pathways given the established link between those who suffer the most from the impact of climate change and pollution, and those who face systemic socio-economic and healthcare inequalities.^26^

Moreover, to enhance the generalisability of the findings, a more comprehensive, national trial and audit should be conducted to determine the efficacy of iOWNA’s support tool across different patient care disciplines and also realise the benefits observed in this audit on a more extensive scale. A national audit would have a role in creating the consensus required to define gold standards for the implementation of digital tools for the management of IBD patients.

## CONCLUSION

The deployment of iOWNA’s digital tool had a significant positive impact on care delivery in IBD clinics at MKUH, reducing unnecessary in-person educational consultation appointments and clinician time spent on information dissemination to patients, contributing to increased PIFU rates and to financial savings for the hospital, as well as improving both clinician and patient experience of the clinics.

The audit’s results underscore the importance of leveraging the digital transformation of care pathways to enhance healthcare efficiency and improve patient care, which will be an essential part of developing healthcare systems that are fit for the future, in which the efficiency of care delivery is improved, waiting lists are cut, patient engagement is boosted, and clinician burn-out reduced.

## Data Availability

All data produced in the present work are contained in the manuscript

